# Strategies to support early- and mid-career cardiovascular researchers to thrive

**DOI:** 10.1101/2022.02.06.22270563

**Authors:** Emma E Thomas, Niamh Chapman, Soraia de Camargo Catapan, Rachel E Climie, Steven G. Wise, Katrina M. Mirabito Colafella, Dean S. Picone, Sally C Inglis, Joanne T.M. Tan, Jason Wu, Lauren Blekkenhorst, Anna C. Calkin, Francine Z. Marques

## Abstract

**Background:** Recent evidence indicates that high numbers of cardiovascular (CV) researchers have considered leaving the research and academic sector due to lack of job security and low funding success. Thus, there is an urgent need to develop solutions to support the retention of early- and mid-career researchers (EMCRs). Here, we aimed to explore the current challenges faced by CV EMCRs, identify solutions to support their career progression and retention, and define a pathway forward to provide a thriving CV EMCR culture in Australia.

**Methods:** Australian CV EMCRs (<15 years post-PhD; n=34) participated in 90-minute online focus groups (n=7) to examine current CV research culture, equity in career progression and solutions (including a timeframe and level of priority) to overcome challenges to career success. Participants were purposefully grouped based on socio-demographic information, including years post-PhD, gender, ethnicity, sexual orientation and caring responsibilities.

**Results:** Participants identified that current metrics only rewarded a narrow set of successes and did not support a collaborative culture. The current appraisal of career disruption in grant applications was identified as inadequate to address underrepresented researchers, such as women and those from culturally-diverse backgrounds. EMCRs proposed 92 solutions aimed at interpersonal, organisational or external levels, with capacity building and equitable opportunities as key focus areas.

**Conclusion:** Pragmatic, cost-effective and implementable opportunities were identified to support the career progression of CV EMCRs to create a more sustainable, equitable and supportive workforce. This information can be used to strategically engage key stakeholders to enable CV EMCRs to thrive.

## Introduction

Cardiovascular (CV) disease remains a major cause of death and disability in Australia.^1^ It is evident that investment in CV research improves the health and wellbeing of people living with CV, with each dollar spent on the sector estimated to result in a 9.8-fold economic benefit.^2^ Given the ageing population in Australia and increasing rates of chronic disease^3^, it will be pivotal to have a strong research sector to continue to drive innovation and evidence-based care. However, a recent survey of Australian CV researchers determined that a lack of job security and limited funding were driving researchers out of the sector.^4^ In fact, a staggering 91% of respondents indicated that they would leave the sector if their position could not be funded in the next few years.^4^ Several gender based issues were also identified in the survey, with female CV researchers being less likely to hold a leadership position and twice as likely to consider leaving the CV sector compared to male researchers.^4^

This workforce survey also highlighted the lack of evidence-based, strategic solutions to support researchers to thrive in CV research.^4^ As early- and mid-career researchers (EMCR) will become the next generation of CV research leaders, actions to ensure their retention are of critical importance to ensure a strong workforce of the future. Thus, EMCR perspectives on how to improve the sector are of paramount importance to strategically develop practical recommendations to guide key stakeholders (e.g. funders, CV research alliances, medical research institutes and universities) to address the challenges faced by CV EMCRs. The present study aimed to qualitatively identify the challenges experienced by CV EMCRs in Australia. Here, we describe our findings and explore solutions to support career progression to increase the retention of EMCRs in the Australian CV research sector.

## Methods

### Study design

Ethics approval was obtained from The University of Queensland (#HREC/2021/HE000419), and informed consent was obtained from all participants. Focus groups with Australian CV EMCRs were conducted according to a specific schedule to examine (i) the current and ideal culture of CV research; (ii) equity in career progression within CV research; and (iii) solutions (including a timeframe and level of priority) to overcome present challenges faced by EMCRs. Consolidated Criteria for Reporting Qualitative Study (COREQ) checklist guided the reporting of findings (Supplementary File – Appendix A).^2^

### Participant recruitment

In April 2021, Australian EMCRs (<15 years post PhD) who worked in the CV research sector within the past five years were invited to participate in a 90-minute online focus group. Participants were recruited via the Australian Cardiovascular Alliance (ACvA; a peak body for CV researchers) member mailing list, social media channels (e.g. Twitter) and through relevant institutes, universities, and CV research networks. Interested participants completed an online expression of interest form which included a link to the participant information sheet, online consent and requested their contact details, socio-demographic information (including years post-PhD, gender, ethnicity, sexual orientation, and caring responsibilities), and interview availability. Participants were purposefully grouped based on their socio-demographic information to facilitate discussion of gender- and diversity-specific issues. The groups included an all-male group, two all-female groups, one with primary caregivers and three mixed groups.

### Data collection

Between May and June 2021, seven focus groups were conducted online via the Zoom (https://zoom.us/) platform. Two female CV researchers (NC, EET) who are experienced in qualitative research, conducted all focus groups. As it is relatively small field some of the participants were known to the facilitators. The full interview guide was piloted with the authorship group and is detailed in the Supplementary File (Appendix B). Focus group sessions were recorded via the Zoom platform and automatically transcribed using the Zoom automated transcription software. The researchers documented reflective notes after each focus group and discussed potential areas that may require further discussion in future groups.

### Data analysis

Recording transcriptions were reviewed for accuracy and de-identified by one researcher (SCC). Content analysis^4^ was used to study both current and ideal CV culture. Thematic analysis, as described by Braun and Clark,^5^ was used to determine the main disadvantaged groups, how they experience disadvantage within the CV sector, and potential solutions to support EMCRs. The proposed solutions were synthesised into level of priority (high >5 votes; medium = 3-5 votes; low <3 votes) based on the number of participants that individually selected the solution as their highest priority (using the Zoom polling function). Participants were also asked to discuss if they thought the solution was able to be implemented within the short (1-2 years), medium (3-5 years) or long term (>5 years).

### Trustworthiness and rigour

According to Guba,^6^ trustworthiness of qualitative research has four constructs: credibility, transferability, dependability and confirmability. Credibility of the study was enhanced by all of the investigators being CV EMCRs. Transferability was improved by including researchers from diverse geographical settings, with a mix of research areas (e.g. basic/discovery, public health, clinical) and career stages. The purposeful grouping of certain participants (e.g. all male, all female, all caregivers) enabled more in-depth discussions around gender-specific issues and challenges of balancing work and family responsibilities. Dependability was ensured by involving an independent researcher (SCC) to perform the first coding pass of the data. Confirmability was then established by three peer-debriefing and consensus meetings, which were conducted during the data analysis stage to discuss the codes and categories and data saturation.

## Results

### Group and participant characteristics

Participant (n=34, mean n=5/group) demographics are provided in Table 1. In brief, 71% of participants were female, 50% were of European decent, and 3% identified as lesbian, gay, bisexual transgender, gender diverse, intersex, queer, asexual or questioning (LGBTIQA+). All Australian states and territories were represented except for the Northern Territory. Most participants were within five years post-PhD (73%), and reported having caregiver responsibilities (71%). A quarter (26%) cared for one or more children under the age of six years.

**Table 1.** Demographics of Australian early- and mid-career researchers who participated in the online focus group discussions

| Position | Categories | Count (%) |
| --- | --- | --- |
| Gender | Female | 24 (71) |
|  | Male | 10 (29) |
| Years post-PhD | 0 – 3 | 15 (44) |
|  | 4 – 5 | 10 (29) |
|  | 6 – 10 | 4 (12) |
|  | >11 | 5 (15) |
| Australian State or Territory of Residence | Victoria | 11 (32) |
|  | New South Wales | 8 (23) |
|  | Queensland | 6 (18) |
|  | South Australia | 3 (9) |
|  | Tasmania | 3 (9) |
|  | Australian Capital Territory | 2 (6) |
|  | Western Australia | 1 (3) |
| Gender identity and sexual orientation | Non-LGBTIQA+ | 28 (82) |
|  | LGBTIQA+ | 1 (3) |
|  | Prefer not to say | 4 (12) |
|  | Did not respond | 1 (3) |
| Ethnic background | European-descendent | 17 (50) |
|  | Non-Caucasian | 11 (32) |
|  | Southeast Asian | 2 (6) |
|  | Other | 1 (3) |
|  | Prefer not to say | 3 (9) |
| Area of research | Clinical | 12 (35) |
|  | Basic/discovery | 8 (24) |
|  | Public health | 4 (12) |
|  | Implementation | 1 (3) |
|  | Other | 9 (26) |
| Family responsibilities | Care for one or more children (including children $\leq 5$ years of age) | 12 (35) |
| | Care for one or more children (all children $>5$ years of age) | 6 (18) |
|  | Other family/caring responsibilities | 6 (18) |
|  | Do not have family/caregiver responsibilities | 9 (26) |
|  | Did not want to say | 1 (3) |

Legend: LGBTIQ+: lesbian, gay, bisexual, transgender, gender diverse, intersex, queer, asexual and questioning.

### Description of the current and ideal CV research culture

#### Current culture

Keywords used by participants to describe the current CV culture are displayed in Figure 1. While many felt there were supportive national collaborations, the most common word across the focus groups was competitive, stemming from limited funding opportunities and low success rates making it “*very difficult to attract funding*”. It was felt that opportunities were disproportionately provided to a few dominant research groups in a “*success begets success”* model. There was also a perception that expectations and workload were constantly increasing, and “*you can never do quite enough*”.

**Figure 1.**
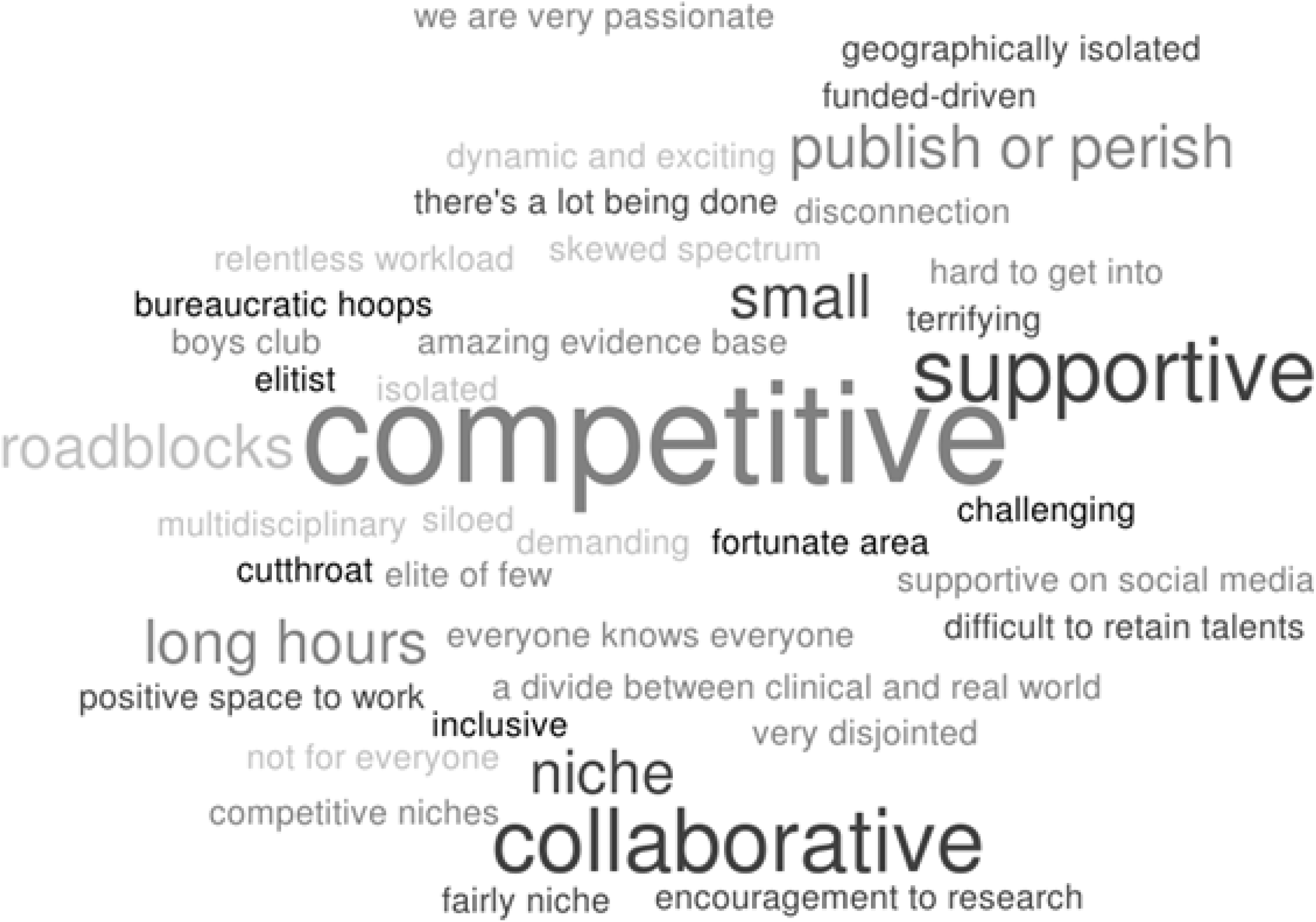
Word cloud of current CV culture according to Australian early- and mid-career researchers that participated in online focus groups. The larger the word, the more frequently it was mentioned by participants.

#### Ideal culture

When asked about the perceived ideal CV culture in Australia, participants envisaged a supportive, collaborative, team-based environment (Figure 2). Mentorship, inclusive leadership and having transparent metrics of success were terms associated with an ideal culture. It was also mentioned that greater recognition and appreciation of EMCRs was required. Moreover, opportunities to *“celebrate success”* would foster a positive work environment.

**Figure 2.**
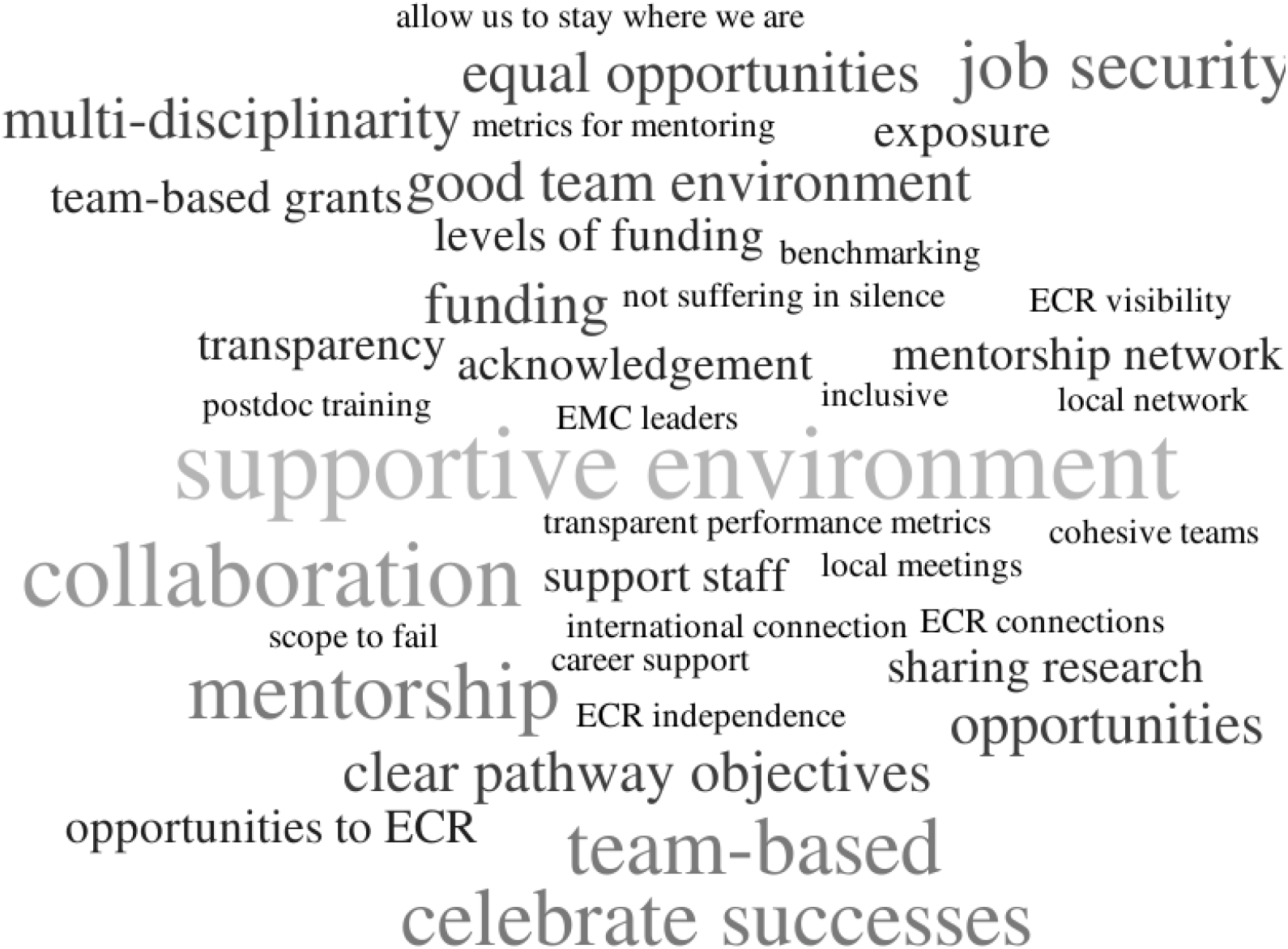
Word cloud of ideal CV culture according to Australian early- and mid-career researchers that participated in online focus. The larger the word, the more frequently it was mentioned by participants.

### Equity and Diversity

Participants identified three key groups that were disadvantaged with regard to career progression in the CV research sector: women, culturally- and linguistically-diverse (CALD) researchers, and researchers based in small research teams (even if situated within large institutions; Table S1). In addition, it was felt that the current CV research workforce does not reflect the diversity of individuals living with CV disease. For example, the lack of Aboriginal and Torres Strait Islander researchers working in the CV research sector was noted. Furthermore, it was felt that funding success did not reflect the health workforce in Australia, with a perception that grants are disproportionately awarded to clinician researchers with medical backgrounds over allied health and nursing researchers.

#### The system favours men

Participants perceived that the CV research sector is male-dominated due to historic and ongoing systemic bias with the current model of success. Participants frequently identified women as being disadvantaged due to unconscious bias, disproportionate caring responsibilities and career disruptions, which limit the capacity to build track record according to current metrics of success such as publications, conference attendance and project leadership. Women described an ongoing tension between family and work commitments and a belief that part-time work would damage their career. The timing of starting a family during the early stages of building a career, in many cases post-PhD, was also reported to have an exponential negative impact on career trajectory due to loss of momentum required to succeed in academia. The pressure to compete with colleagues without career disruptions was further challenged by the perception that criteria of the impact of *career disruptions* within grant applications is not uniformly applied and does not quantitatively reflect the ‘real life’ impact beyond the specifically defined period of leave *per se*. Illustrative quotes are shown in Table S1.

#### Culturally- and linguistically-diverse (CALD) researchers face additional disadvantages

Non-Australian citizens experience additional challenges as most grant and fellowship criteria require applicants to be Australian permanent residents or citizens. Thus, the lack of eligibility to apply for, and secure such funding opportunities can have a significant impact on career progression. In addition, they may experience career disruptions to focus on visa applications or other activities to ensure their stay in the country. Concerningly, instances were described where people felt compelled to do extra work for their team leader to ensure the continuation of their contract and/or visa status. Researchers for whom English is a second language also face the additional challenge of communication barriers and can take longer to produce manuscripts, grants and presentations, reducing overall productivity.

#### Isolation: Small cogs in small wheels

Some researchers experienced a sense of isolation and invisibility caused by geographical distance, lack of good mentorship, being in a small institution, or being part of a group that did not have prominent visibility at a national level. It was felt that this isolation led to fewer opportunities and outputs such as group-based publications. Some described themselves as being *“an island”* rather than *“part of a bigger machine*”.

### Solutions and Recommendations

In total, 92 solutions were generated, which were consolidated into 29 solutions grouped under the themes *build capacity to support success* and *opportunities for all* (Table 2).

**Table 2.** Solutions generated by the focus groups.

| Suggested idea or solution | Timeline | Priority |
| --- | --- | --- |
| **Build capacity to support success** |  |  |
| *Support collaboration between researchers* |  |  |
| Develop a public database/website of CV researchers profiling EMCRs including areas of interest, skills, and collaborations they are seeking. Increase visibility by providing researcher highlights | Short-term | High |
| Provide seed-funding for EMCRs to encourage the development of collaborations | Medium-term | High |
| Support the development and growth of CV networks (e.g. QCVRN, NSW-CVRN) to advocate for CV researchers | Long-term | High |
| *Support collaboration between researchers and clinicians* |  |  |
| Encourage local networks to develop annual meetings where clinicians and researchers pitch research ideas | Short-term | Medium |
| Develop funding schemes that require clinician-researcher investigator teams | Medium-term | Medium |
| Co-localise clinicians and researchers | Long-term | Medium |
| Develop EMCR Industry fellowships to enable opportunities to work with industry and clinical partners | Long-term | Medium |
| *Support collaboration between researchers and patients* |  |  |
| Develop programs to promote connections with patients and skills in patient engagement/co-led research | Short-term | Low |
| Enhanced networks with clinicians/patients to improve research translation | Medium-term | Low |
| Develop an Australia-wide patient group | Long-term | Low |
| *Provide training during early post-doctoral training* |  |  |
| External training to promote leadership and management skills | Short-term | Medium |
| Internal (institution-specific) training on non-research skills (e.g., finance, grant administration, recruitment of staff) | Medium-term | Medium |
| *Enhance and incentivise mentorship and sponsorship* |  |  |
| Provide mentoring opportunities, especially during periods of transition, with training to support good mentorship | Short-term | Medium |
| Group-based mentorship – expectation of grant success that you partner with and support another (e.g. smaller) group<br>(e.g. part of funding used to extend broader collaborations) | Medium-term | Medium |
| Provide access to career coaching and strategic coaching | Short-term | Medium |
| Quantify, record and recognise how people have supported others to succeed (e.g. in performance appraisals, grants) | Long-term | Medium |
| Embed mentorship into metrics of success (e.g. be required to state proof of mentorship and how it has helped others) | Long-term | Medium |
| *Support EMCRs in funding schemes* |  |  |
| Develop opportunities for EMCRs to lead aspects of grants (e.g. co-CIA with more senior researchers) and/or incentivise senior researchers to have EMCRs as CIs. | Short-term | High |
| Simplify the application process to reduce unnecessary time spent on applications by offering a short expression of interest with invited submission for shortlisted applications, centralise the grant application process, and standardise track record statements/sections. | Short-term | Medium |
| Provide clear performance metrics for each career stage with clear benchmarking nationwide to facilitate comparison and minimum transparent expectations across career stages and areas of research (e.g. basic/discovery versus clinical). Such metrics should be used to support funding schemes to set eligibility based on set minimum performance standards (i.e. sufficient track record across the project team to be feasible), while ranking would be based specifically on project or idea merit. | Medium-term | Medium |
| Facilitate a team-based approach that recognises collaboration (e.g. CI team reflects contribution with each CI leading a component of the research), values different skillsets and supports researchers to maintain a position rather than progressing towards to professor or group leader. | Long-term | Medium |

| Equitable opportunities |  |  |
| --- | --- | --- |
| *Parental and caring responsibilities* |  |  |
| Support conference attendance by providing on-site childcare, and financial support for family member/additional caregiver to attend and continue options for virtual or remote access. | Short-term | High |
| Maintain momentum during/immediately after leave periods including research assistants to progress projects during and on return from leave periods, a clear and low-burden process to extend funding for leave periods, on-site or support for childcare, institutionally standardised dedicated time for writing. | Medium-Long-term | High |
| Change expectations and societal norms surrounding parental duties by extending and normalising leave periods for both parents, including parental responsibilities on funding applications (even if workload did not decrease), support flexible and part-time working arrangements for all parents/individuals with caring responsibilities. | Long-term | High |
| *Supporting underrepresented groups* |  |  |
| Provide greater support for preparing funding applications through mentorship, coaching, and writing support for women and CALD researchers. | Short-term | Medium |
| Remove identifiable information from grants and offer specific schemes or allocate a specific amount of funding for | Long-term | Medium |
| underrepresented groups including women and/or international researchers. |  |  |
| *Increase visibility of EMCRs* |  |  |
| EMCR presentation opportunities such as: ‘meet the author’ event across institutions, invited senior and EMCR presentations between organisations, shared profiles facilitating introductions, disseminate skills that people/teams are looking for through social media or databases. | Short-term | Low |
| Enable EMCRs from smaller/regional groups to work with larger groups via targeted collaboration grants | Medium-term | Low |
| Develop ‘sister laboratories’, i.e. partnerships between small/large groups across CV institutions/groups<br>Facilitate ways for smaller groups to access equipment from larger groups | Long-term | Low |

**Legend:** CALD, culturally- and linguistically-diverse; CI, chief investigator; CIA, chief investigator A; CV, cardiovascular; NSW-CVRN, New South Wales Cardiovascular Research Network; EMCRs, early- and mid-career researchers; QCVRN, Queensland Cardiovascular Research Network.

#### Build capacity to support success

Participants identified multiple ways to build capacity among EMCRs to support career progression. The key areas to build capacity were: 1) support collaborations between researchers, clinicians and consumers; 2) provide training for skills required to succeed beyond those included in scientific research training, such as finance, people management, science communication; 3) provide mentoring and/or coaching alongside rewarding effective mentorship to incentivise the practice, particularly among successful and more senior researchers; 4) increase the visibility of EMCRs to facilitate networking, collaboration and establishing independence and 5) increase support for EMCRs in funding schemes with clear metrics for success, facilitating a team-based approach and rewarding teams with an EMCR chief investigator(s).

#### Equitable opportunities

There were three key areas that solutions for equitable opportunities centred around: those with parental/caring responsibilities, underrepresented researchers and greater visibility for EMCRs (Table 2). Solutions related to parental and caring responsibilities that could be easily implemented at little or no cost included: 1) avoidance of school holidays for grant and fellowship deadlines and 2) addressing the expectation that you need to be responsive to work demands during weekends and periods of leave; 3) providing dedicated time for writing; 4) acknowledgement of parental responsibilities in funding applications and 5) providing virtual or remote access to seminars and conferences. Stipends to support a carer for conference travel (with access to onsite childcare facilities) and to support a research assistant for individuals on parental leave or in their transition to returning to work to facilitate the continuation of their research projects were also suggested. However, it was discussed that, in the long-term, a greater cultural shift was required at the policy and societal level, to enable families to divide parental leave arrangements more evenly.

Solutions to support underrepresented groups included mentorship and coaching, with a particular focus on writing and securing funding, as well as the specific allocation of funds for CALD and female researchers. Solutions to increase the visibility of EMCRs included skills databases, targeted collaboration grants as well as partnerships and networking between research groups and institutes.

## Discussion

Using qualitative research methods, this study has identified priority solutions to improve the culture in the CV research sector that will support Australian EMCRs. The proposed solutions from EMCRs include better support and opportunities for collaboration, training, mentorship, new metrics of success, and the removal of barriers to improve participation of underrepresented groups. Importantly, the findings from this study are targeted towards strategic implementation as solutions were prioritised with a short-, medium- or long-term timeline indicated.

### Key findings about culture

The ideal work culture that participants valued most was a cohesive, collegial, and collaborative environment that provides support and mentorship. Indeed, studies have shown that when individuals, especially those from minority groups, feel valued, supported, have a sense of ‘belonging’ and had clear departmental/Faculty expectations, they were more likely to be productive and have increased publication outputs.^7, 8^ To enable the sector to move towards team-based approaches, funding needs to be directed to teams, with varying skills and levels of experience, rather than at an individual level.

### Building capacity to support success

One of the limitations for EMCRs was a lack of visibility to establish peer collaborations. The development of databases where skills and resources could be shared would increase visibility and assist in the development of whole-of-pipeline collaborations to support cross-sector engagement. The second limitation identified was access to seed funding to support the development of early collaborations and ideas. Australia has several successful examples of the power of seed funding to generate impactful CV research. Seed funding from the National Heart Foundation of Australia was essential for the initial feasibility studies of the quadpill, now confirmed to achieve better blood pressure control than monotherapy in a landmark clinical trial.^9^ Another example includes the discovery of new mechanisms of blood pressure regulation in pre-clinical models,^10^ which was supported by seed funding from the Foundation for High Blood Pressure Research, and now has led to a randomised clinical trial.^11^ Existing funding schemes could be redesigned to require EMCR leadership and allow recognition of EMCRs as main/equal chief investigators. Moreover, EMCRs urged funding bodies to simplify funding applications, such as by offering two stage applications (expression of interest for first stage, full application for invited applicants at second stage), which have been shown to be more cost- and time-effective than one stage applications.^12^ State-based CV research networks, such as those developed in New South Wales, Queensland and Western Australia, could have a key role advocating these and other changes for researchers; a powerful approach if done in a harmonised way, and aligned with national advocacy as led by the ACvA.

Cross-sector engagement, however, is not limited to peer-collaboration; it also encompasses researcher-clinician, researcher-consumer, and industry partnerships. EMCRs highlighted that whilst they appreciate the importance of researcher-clinician collaborations, these were challenging to establish. Indeed, barriers to researcher-clinician collaborations are well documented.^13^ A change in culture, however, is possible with substantial two-way opportunities for training and career development available.^14^ Suggestions to minimise the silos segregating researchers and clinicians included providing a favourable environment for success such as the co-location of clinicians and researchers. There are many successful examples of co-location in Australia, such as the Baker Heart and Diabetes Institute/Alfred Hospital, the Murdoch Children’s Research Institute/Royal Children’s Hospital, and the Victor Chang Cardiac Research Institute/St Vincent’s Hospital Sydney, amongst others. Moreover, existing funding schemes could be restructured to incentivise teams of clinicians and researchers. Similarly, meaningful engagement with consumers was highlighted as important for research translation but difficult to establish by single individuals. Institutional or non-government organisations could facilitate connections with consumers, with a long-term goal being the development of an Australia-wide consumer group. In addition, more efficient alignment of research themes with the health challenges through strategic partnership between government and health could lead to more efficient investment in research, allowing researchers to focus on solving the key problems with impact. Given the direct health care costs of CV disease per year (>$10 billion),^15^ shifting the dial on this through whole of pipeline research and implementation solutions could lead to a more sustainable and thriving ecosystem.

A major skill EMCRs need to develop prior to consumer engagement is effective science communication. A successful example of how this can be achieved includes the ACvA CV Champions Program, which provided science communication skills training and tools to 51 researchers and clinicians across Australia over a 12-month period during 2020-21.^16^ Finally, building strong partnerships between academia and industry is essential, and has been one of the major focuses of the ACvA.^17^ This could be strengthened by the development of CV industry fellowships as suggested by our participants, such as the Researcher Exchange and Development within Industry (REDI) fellowships,^18^ and a dedicated industry-academia cross sector mentoring scheme.

Indeed, another important aspect to build capacity is mentoring, particularly during periods of transition, such as from PhD to post-doctoral training or return from parental leave. There is a plethora of evidence regarding the importance and the power of strategic mentoring for career development overall^19^ and in cardiology specifically.^20^ Mentoring also constitutes an important aspect to support EMCRs during and post the SARS-CoV-2 pandemic.^21, 22^ Yet, several EMCRs highlighted that they did not have mentors. Developing and engaging EMCRs in mentoring schemes, such as the ACvA Cross-Sector Mentoring Scheme, aimed at connecting researchers, clinicians, industry and policymakers,^16^ can fulfil this demand. Our participants also emphasised the need for mentoring to be recognised as an official metric of success and to be part of performance appraisals, embedding its importance in career progression. In this respect, the ACvA have recognised the importance of mentoring through a dedicated national award for mentorship. Finally, EMCRs highlighted the lack of leadership, management and financial skills, among others, which are essential for career progression in CV research. A solution was to provide specific training and support in these areas at the early-career stage.

### Equitable opportunities

The benefits of a diverse research sector have been demonstrated across many facets, from greater scientific innovation and enhanced public trust, to more highly cited publications with greater impact.^23, 24^ This highlights the need to remove the barriers and biases’ faced by underrepresented groups to ensure that we have a diverse CV research sector that can most effectively address the burden of CV disease. EMCRs identified several strategies to address the ‘barriers and bias’ faced by underrepresented researchers including CALD individuals and those with parental and caring responsibilities.

A number of solutions were presented around funding applications, which could be incorporated at little cost, but would have significant impact on the opportunities for career progression for a diverse range of individuals. These included the incorporation of metrics that fairly and transparently evaluate researchers from different career pathways, fields of research, backgrounds, abilities, work and life experiences. Metrics should also incorporate internal and external contributions, such as mentoring, which are critical to the workplace and sector, particularly given that women take on a disproportionate amount of ‘service’ roles.^25^ For example, the National Health and Medical Research Council (NHMRC) Investigator Grant Scheme now incorporates research mentoring and institutional leadership as two of the four components that make up the leadership section of the application.^26^ They now also recommend the use of gender neutral language, which is important given the recognised impact of gendered language on grant success favouring males.^27^ However, men continue to disproportionally receive more funding.^28^ This could be addressed by funding bodies allocating a similar percentage to men and women and other minorities. It was further suggested that funding bodies provide a clear and low-burden process for extending funding for periods of leave, which is critical given the time-dependent nature of research. Moreover, clear criteria and training to provide consistency regarding the assessment of *career disruptions* and *relative to opportunity* sections of funding applications was discussed. Indeed, whilst granting bodies allow for the addition of track record for a period of time commensurate with the leave taken, in the case of parental leave, this by no means reflects the impact of parental responsibility once an individual returns to work. EMCRs suggested the inclusion of a section within grant application for parental responsibilities to reflect this impact. As a comparison, the Australian Research Council (ARC), NHMRC sister’s research council, provides two years of career interruption (inclusive of parental leave) for a primary carer regardless of the time formally taken for parental leave.^29^

Grants to support a carer to allow a researcher to bring young children with them when attending a conference were suggested. Given that conference presentations are a key metric for assessment of track record as well as providing networking and collaborative opportunities, such grants could have a significant impact on career development, particularly at a time when career trajectory is impacted. Similarly, grants that support a research assistant whilst on leave will facilitate maintenance of some level of momentum in an individual’s research program whilst on leave and/or in the period when they are transitioning back to work. One example of such a program is the Advance Queensland Women’s Research Assistance Program.^30^ Across the sector other funding opportunities have been developed such as the Franklin Women Travel Scholarship^31^ and the Susan Alberti Women in Research Award.^32^ Funding policies to allow CALD researchers to apply also need to be considered, given the importance of securing funding for track record and career progression. Equitable opportunity to secure grants is particularly critical given the competitive, time-dependent nature of research and was identified as a high priority action in the discussion of research culture.

Lastly, flexibility was presented as a solution to address inequities across the CV research sector. Flexible work hours and access to virtual seminars and conferences will advantage not only those individuals with parental and caring responsibilities, but also researchers that travel long distances to workplaces or are geographically isolated, and in the case of virtual conferences, those that lack the travel funds to attend conferences. These opportunities also come at low cost, yet have a significant impact on inclusivity and address many of the barriers faced by underrepresented groups. The way in which we have responded to the SARS-CoV-2 pandemic, with many Australians living through lockdowns for much of 2020-21, has demonstrated that flexible and remote arrangements are a feasible, effective and inclusive strategy that could readily be continued.

### Strengths and limitations

This study was strengthened by the purposive sampling approach which enabled a broad diversity of EMCRs viewpoints with respect to geographical location, research field, carer responsibility and ethnic background within the CV research sector in Australia. This approach ensured that we identified both issues affecting these groups and their perspective on actions and solutions to address them. The solutions generated with and for EMCRs are, therefore, highly relevant to the sector and provide a list of actions to present to stakeholders, that includes several practical and cost-effective actions that can be implemented immediately. Unfortunately, we were unable to recruit any researchers that identified as having a disability or Aboriginal or Torres Strait Islander. We acknowledge that while our findings may have broader relevance to other settings and sectors, the focus of this research was the Australian CV research sector. Lastly, due to COVID-19 restrictions, the focus groups were conducted online. We acknowledge that more in-depth discussions may have occurred in person. However, a strength of the online format is that it did enable a greater diversity of participants to take part, particularly from underrepresented and geographically diverse locations, given that no travel was required.

### Future Directions

Collaboration with key organisations such as funding bodies, institutes and universities will be crucial to facilitate implementation of these solutions. To drive change forward, action needs to be measured and assessed, to ensure accountability such as through developing key performance indicators. Further, EMCRs need representation on panels related to grant development and key decisions that will impact their careers.

## Conclusion

We have identified a raft of solutions with and for EMCRs to support a sustainable and connected CV research community. The status quo will not do. Today’s EMCRs and tomorrow’s future leaders are looking for inclusive and supportive workplaces and are wishing to transform competitive workplaces to ones of collaboration. This information can be used to strategically engage key stakeholders. To enable change to occur, action will be required across multiple levels from EMCR-led initiatives and representation to the organisational and policy level, with the support of senior leadership.

## Data Availability

All data produced in the present study are available upon reasonable request to the authors

## Acknowledgements

We thank the Australian Cardiovascular Alliance (ACvA) Board for their support. We acknowledge the supporting roles of ACvA President, Professor Gemma Figtree, ACvA Chief Executive Officer, Kerry Doyle and ACvA Project Officer, Meng-Ping Hsu, in the facilitation of this initiative. We acknowledge that the authors (with exception of S.C.C.) were members of the ACvA Emerging Leaders Committee at the time this research was conducted. We also acknowledge other members of the Emerging Leaders Committee including Lauren Blekkenhorst.

