## Supplemental Data 1 for "Strategies to support early- and mid-career cardiovascular researchers to thrive"

### **Appendix A – COREQ checklist**

| **No. Item** | **Guide questions/description** | **Reported on page** |
| --- | --- | --- |
| **Domain 1: Research team and reﬂexivity** | | |
| *Personal Characteristics* | | |
| 1. Inter viewer/facilitator | Which author/s conducted the interview or focus group? | 6 |
| 2. Credentials | What were the researcher’s credentials? E.g. PhD, MD | 6 / Title pg |
| 3. Occupation | What was their occupation at the time of the study? | 6 / 7 |
| 4. Gender | Was the researcher male or female? | 6 |
| 5. Experience and training | What experience or training did the researcher have? | 6 |
| *Relationship with participants* | | |
| 6. Relationship established | Was a relationship established prior to study commencement? | 6 |
| 7. Participant knowledge of the interviewer | What did the participants know about the researcher? e.g. personal goals, reasons for doing the research | App B |
| 8. Interviewer characteristics | What characteristics were reported about the inter viewer/facilitator? e.g. Bias, assumptions, reasons and interests in the research topic | App B: Interview guide |
| **Domain 2: study design** | | |
| *Theoretical framework* | | |
| 9. Methodological orientation and Theory | What methodological orientation was stated to underpin the study? e.g. grounded theory, discourse analysis, ethnography, phenomenology, content analysis | 7 |
| *Participant selection* | | |
| 10. Sampling | How were participants selected? e.g. purposive, convenience, consecutive, snowball | 6 |
| 11. Method of approach | How were participants approached? e.g. face-to-face, telephone, mail, email | 6 |
| 12. Sample size | How many participants were in the study? | 7 |
| 13. Non-participation | How many people refused to participate or dropped out? Reasons? | NR |
| *Setting* | | |
| 14. Setting of data collection | Where was the data collected? e.g. home, clinic, workplace | 6 |
| 15. Presence of non-participants | Was anyone else present besides the participants and researchers? | NR |
| 16. Description of sample | What are the important characteristics of the sample? e.g. demographic data, date | 7/8 Table 1 |
| *Data collection* | | |
| 17. Interview guide | Were questions, prompts, guides provided by the authors? Was it pilot tested? | App B |
| 18. Repeat interviews | Were repeat inter views carried out? If yes, how many? | No |
| 19. Audio/visual recording | Did the research use audio or visual recording to collect the data? | Yes, pg 6 |
| 20. Field notes | Were ﬁeld notes made during and/or after the interview or focus group? | Yes. Pg 6 |
| 21. Duration | What was the duration of the inter views or focus group? | 6. |
| 22. Data saturation | Was data saturation discussed? | NR |
| 23. Transcripts returned | Were transcripts returned to participants for comment and/or correction? | No |
| **Domain 3: analysis and ﬁndings** | | |
| *Data analysis* | | |
| 24. Number of data coders | How many data coders coded the data? | 7 |
| 25. Description of the coding tree | Did authors provide a description of the coding tree? | NR |
| 26. Derivation of themes | Were themes identiﬁed in advance or derived from the data? | No |
| 27. Software | What software, if applicable, was used to manage the data? | NR |
| 28. Participant checking | Did participants provide feedback on the ﬁndings? | No |
| *Reporting* |  |  |
| 29. Quotations presented | Were participant quotations presented to illustrate the themes/ﬁndings? Was each quotation identiﬁed? e.g. participant number | Table S1 |
| 30. Data and ﬁndings consistent | Was there consistency between the data presented and the ﬁndings? | Yes |
| 31. Clarity of major themes | Were major themes clearly presented in the ﬁndings? | 8-11 |
| 32. Clarity of minor themes | Is there a description of diverse cases or discussion of minor themes? | 8-11 |

### **Appendix B – Interview guide**

**[Introductions]**

**Background –** previous ACVA survey and work and aim to develop solutions to enable CV researchers to thrive in the sector.

**Research culture** – describe current culture and identify areas for improvement.

Prompts:

a. What comes to mind, in a word or two, to describe CV research culture in Australia?
b. What would the ideal culture look like for you?
c. How could we move towards that ideal culture?
d. What is already working well and how can we build on that?

**Diversity and equity** – describe specific diversity, equity and inclusion challenges and identify. Highlight survey findings.

Prompts:

a. Do you think any minority or underrepresented groups experience any specific challenges in CV research that others may not? If so, what are they?
b. Why do you think these groups experience such challenges?
c. What could be done to support these groups in CV research?

**CV research solutions**.

ACTIVITY:

i. Spend 1 minute and write down all the solutions associated with identified (already identified solutions highlighted)
ii. Discuss solutions and rank into short, medium, long-term
iii. Prioritise solutions using polling function

***Thank you for your time***

**Table S1.** Illustrative quotes from participants regarding equity and diversity.

| **Theme** | **Sub-theme** | **Exemplar Quotes** |
| --- | --- | --- |
| **The system favours men** | *Clinical CV research is male dominated* | And as a male I would have to acknowledge that I see challenges for female colleagues going through clinical cardiology research as well. (…) They're all challenging things to rectify but it's historically a male dominated workforce - the clinical cardiology world. |
|  |  | I was actually working while I was in labor - it was not planned that way, but my daughter came too early. So, I was at the office but it was painful (...) they told me the first one [child] actually takes 20 hours sometimes. [So I had to] wrap up what I was doing, take photos so (…) they know what to do next. But I don't think that's a good model. I've been telling my students, because I just feel like this is just too much pressure. Why was I doing it? It was because I felt challenged. Well, actually, I feel threatened a little bit because most of my peers are actually guys. |
|  | *Women experience disproportionately greater career disruption* | I think, as a female in academia and research, you definitely have a hard journey, especially when you're at the point where you're having children (…), and maybe you don't want to work full time for a while. I think women definitely have more career disruptions. |
|  | *Parental and caring responsibilities have long-term impacts* | Because what is career disruption about? So, it affects your grant applications, it affects your publications, it affects your business to grow, your ability to go to conference overseas, presentations... So, if there could be some sort of score for that could then be used as a leveller you know, like a weighting or something like that. |
|  | *Grant success is lower in female-dominated professions (e.g. nursing)* | We did some analysis last year around Investigator Grant outcomes and nursing only received 3% of the NHMRC Investigator Grants, but we're 80% of the health workforce in Australia and 90% are women. |
|  | *Success begets success* | When you compare to a man, who (…) during those two years you were absent (…) got a Nature paper. They'll probably still give it to the man, the Nature paper. It's always like people who are very successful they just going to keep succeeding, because the people that haven't had any success yet, it's really hard to open a new door, especially (…) if they're already a minority or women like we haven't been able to have any success it's hard for us to get your foot in the door. |
| **Culturally- and linguistically-diverse researchers face additional disadvantages** | *International researchers are ineligible to apply for multiple funding schemes* | As soon as you've come from an international setting, applying for funding is not an easy thing to do. So often you don't meet the criteria for fellowships and it makes it really hard to progress. If you can't apply for any even small grants, then it becomes a bit of a stopping point for people. |
|  | *Additional requirements to maintain residency status* | I’m taking my case as an example that I do have some disruption, because I needed to sort out my permanent residency application. So I need to do some other work, to study a different degree to stay here, while doing research. (…) Everyone has different situations that can impact their career or their track record. |
|  | *Additionally challenge of writing and verbal communication skills* | I come from non-English speaking country, so it took me a while to be able to get up there and present (..). I don't say neglect, but we need to encourage a little bit more maybe because I’m sure everyone has a brilliant mind, but the way we communicate - it took me a while. |
| **Isolation: Small cogs in small wheels** | *Lack of visibility when not part of a large group* | Of course if you work in a big collaborative group, you have more opportunity, this was the paper, even if you're in a middle [author], but it still justify your work somewhere in there while I came from a very small really niche group. I found it really hard at the beginning, because you can only publish so much in the year. So I think that's also important looking at (…) the size of the group that you are at. |
|  | *Geographic isolation* | I am in a bit of an island this year in terms of where I'm at. |
|  | *Lack of mentoring and support* | And it's also about knowing who to approach. If you don't have the right mentorship that is a huge disadvantage in terms of knowing which areas are good to approach in order to be able to carry out your dream projects. |
